# Frailty and its impact on adverse outcomes in older patients with cancer in Vietnam

**DOI:** 10.1101/2025.03.12.25323826

**Authors:** Tan Van Nguyen, Quyen Ho Hong Tran, Erkihun Amsalu, Trinh Thi Kim Ngo, Thanh Dinh Le, Ying Zhang, Mark Woodward, Tu Ngoc Nguyen

**Author notes:** Corresponding author: Dr Tu Nguyen, The George Institute for Global Health, Sydney, New South Wales, Australia.

## Abstract

**Aims:** This study aimed to quantify the prevalence of frailty, and investigate the impact of frailty on adverse outcomes, in older patients with cancer in Vietnam.

**Methods:** A prospective, observational study was conducted in adults aged 65 or above with cancer who attended the outpatient clinics of two urban hospitals in Vietnam from September 2023 to May 2024. Frailty was defined by the Carolina Frailty Index (CFI) and participants with a CFI >0.35 were identified as frail. All participants were followed up for 3 months after discharge, recording falls, all-cause hospitalization, and all-cause mortality.

**Results:** There were 379 participants (mean age 72.3 years, 48.5% female). The prevalence of frailty was 26.6% (95%CI 22.2% - 31.0%), highest in participants with stomach cancer (35.7%) and lung cancer (33.9%). Participants with advanced stages of cancer had a significantly higher prevalence of frailty: 39.3% in stage 4, 21.7% in stage 3, compared to 18.1% in stage 2 and 13.0% in stage 1. During the follow up, 19.0% of the participants had a fall (44.4% in the frail vs. 9.7% in the non-frail, p<0.001), 33.4% were admitted to hospitals (42.2% in the frail vs. 30.1% in the non-frail, p=0.026). The mortality rate was 1.9% (5.1% in the frail vs. 0.7% in the non-frail, p=0.017). Odds ratios were 7.48 (95%CI 4.24 – 13.40, p<0.001) for falls, 1.71 (95%CI 1.06 – 2.75, p=0.027) for all-cause hospitalization, and 7.10 (95%CI 1.36 – 37.22, p=0.020) for all-cause mortality.

**Conclusion:** Frailty was observed in over a quarter of the participants, with the highest prevalence among those with stomach and lung cancer. Frailty significantly increased the odds of falls, hospitalization, and mortality in three months post-discharge. Further research is needed to gain a better understanding of the impact of frailty on adverse outcomes, and the quality of life for older adults with cancer in Vietnam.

## Introduction

Globally, a significant proportion of adults diagnosed with cancer are older individuals (aged 65 years or above), contributing to approximately 80% of the annual mortality rates associated with cancer.^1^ In older adults with cancer, frailty is common and is of particular importance for this population.^2^ The treatment and follow up for older adults with cancer present significant challenges due to their complex health needs and the presence of multiple chronic health conditions, as well as geriatric syndromes such as frailty.^2^ A systematic review in 2015 of 20 studies (n=2916 participants) from 13 countries (Australia, Austria, Belgium, Canada, France, Germany, Italy, Japan, Norway, Spain, Singapore, the Netherlands, and USA) revealed that the prevalence of frailty in patients with cancer was 42% (range 6%– 86%).^3^ Frailty was independently associated with increased all-cause mortality, increased risk of postoperative mortality, and cancer treatment complications.^2–5^ These highlight the crucial role of frailty assessment in making decisions regarding the management of older patients with cancer.

In Vietnam, it is projected that the burden of cancer incidence will increase significantly in the two largest populated cities in Vietnam (Ho Chi Minh City and Hanoi), with the number of cancer cases being predicted to double from 2013 to 2025 for most types of cancers.^6^ Lung, colorectum, breast and thyroid, and liver cancer represent approximately 67% of the overall cancer burden.^6^ Data from the ASEAN CosTs In ONcology (ACTION) study showed that this is an under-resourced issue, with over 75% of patients with cancer in Vietnam and other Southeast Asian countries experiencing death or family financial catastrophe within one year of cancer incidence.^7^ With a 17% growth in the overall population, and an increasingly aging population in Vietnam, the burden of cancer incidence will increase sharply over the next decades. The prevalence of frailty among older Vietnamese people is also high, with community-based studies reporting rates between 11% and 22% and hospital-based studies indicating frailty prevalence of up to 55%.^8–16^ However, geriatric oncology is an under-researched area, and there is limited evidence of frailty in older adults with cancer in Vietnam. Understanding the burden of frailty in this population is crucial, as it significantly influences both their treatment outcomes and quality of life.

Therefore, this study aimed to quantify the prevalence of frailty and investigate the impact of frailty on adverse outcomes, in older patients with cancer in Vietnam.

## Methods

### Study populations

This prospective, observational study was conducted at the outpatient clinics of two major hospitals in Vietnam (Thong Nhat Hospital in Ho Chi Minh City and Nguyen Trai Hospital) from September 2023 to May 2024. Consecutive patients aged ≥ 65 diagnosed with cancer who visited the clinics during the study period were recruited. The exclusion criteria were: (1) refusal to participate in the study, (2) inability to obtain consent, (3) severe visual or hearing impairment that affected the patient’s ability to answer the study questions.

The study was approved by the Ethics Committee of the University of Medicine and Pharmacy at Ho Chi Minh City (Reference Number 676/HDDD-DHYD). Informed consent was obtained from all participants. This study was conducted in accordance with the Declaration of Helsinki.

### Data collection

Data were collected from patient interviews and medical records. Information obtained included demographic characteristics, lifestyles, height, weight, medical history, duration of having a cancer diagnosis, cancer types, cancer stages, cancer treatment, frailty, and comorbidities. Body mass index (BMI) was calculated from measured weight and height.

Smoking was defined as current smoking (yes/no). Regular exercise was defined as doing exercise at least five days per week (yes/no). Polypharmacy was defined as using 5 or more medications on a daily basis (yes/no).^17^

Comorbidities were documented from the medical records based on a predefined list, including hypertension, coronary heart disease, heart failure, stroke, dyslipidemia, diabetes, chronic kidney disease, osteoarthritis, gastroesophageal reflux disease (GERD), musculoskeletal pain, sleep disorder, depression, and anorexia. In addition, cognitive function was assessed using a 6-item Blessed Orientation-Memory-Concentration (BOMC) test, and a BOMC score ≥5 was defined as cognitive decline.^18^ Anemia was defined as a serum hemoglobin concentration of less than 12 g/dL for women and less than 13 g/dL for men.^19^ Depression was defined using the Geriatric Depression Scale (GDS), and a GDS score ≥5 indicated depression.^20^ The overall comorbidity burden was also assessed using the Charlson Comorbidity Index.^21^

Nutritional status was assessed using the Mini Nutritional Assessment Short Form (MNA-SF), and participants with a score ≤7/14 were identified as having malnutrition.^22^ Risk of falls was assessed using the Stopping Elderly Accidents, Deaths, and Injuries (STEADI) questionnaire.^23^ Activities of daily living (ADL), and instrumental activities of daily living (IADL) were assessed and participants with an ADL score <6 were identified as having ADL impairment, while those with an IADL score <8 were identified as having IADL impairment.

Frailty was defined by the Carolina Frailty Index (CFI).^25^ The CFI was constructed from 36 variables, including instrumental activities of daily living, self-reported health, physical function, comorbidities, number of daily medications, vision, hearing, nutrition, mental health, social activity, and cognition. The CFI scores range from 0 to 1, with higher scores indicating greater levels of frailty. Participants with a CFI >0.35 were identified as frail. ^25^

All participants were followed up for 3 months after discharge, recording adverse outcomes including falls, all-cause hospitalization, and all-cause mortality.

### Sample size calculation

We chose the sample size to be sufficient to estimate the percentage of participants with frailty in cancer patients with an error, taken to be the width of the 95% confidence interval, of no more than ±5%. To compute this requires an estimate of the expected percentage frail, which we took to be 42% based on a systematic review of frailty in patients with cancer worldwide in 2015. ^3^ The resultant sample size requirement was 374.

### Statistical analysis

Continuous variables are presented as means and standard deviation (SD), and categorical variables as frequencies and percentages. Comparisons between frail and non-frail participants were conducted using the chi-square test or Fisher’s exact test for categorical variables and Student’s t-test or Mann-Whitney test for continuous variables.

To quantify the association between frailty and the adverse outcomes of interest (falls, all-cause hospitalization and all-cause mortality), multivariable logistic regression models were applied, adjusting for age, sex, and cancer stages. In addition, we conducted sensitivity analyses, fitting the adjusted models for covariates with p-values <0.05 or <0.25 from univariable analyses (Supplementary Tables 1 and 2). We also conducted subgroup analysis by sex. The results are presented as odds ratios (OR) and 95% confidence intervals (CI) plus women to men ratios of ORs (RORs) with CIs. All statistical tests were two-sided and a p value <0.05 was considered to confer statistical significance.

## Results

We obtained data from 379 participants. They had a mean age of 72.3 (SD 6.0) years and 48.5% were female. The prevalence of frailty was 26.6% (95%CI 22.2% - 31.0%). Frailty prevalence differed among participants with different types of cancer: 35.7% in participants with stomach cancer, 33.9% in participants with lung cancer, 31.3% in participants with prostate cancer, 24.8% in participants with colorectal cancer, 18.1% in participants with breast cancer, 9.1% in participants with liver cancer, and 30.4% in participants with other types of cancer (p=0.226) (Figure 1). Participants with advanced stages of cancer had a significantly higher prevalence of frailty: 39.3% in stage 4, 21.7% in stage 3, 18.1% in stage 2 and 13.0% in stage 1 (p<0.001, Figure 2).

**Figure 1.**
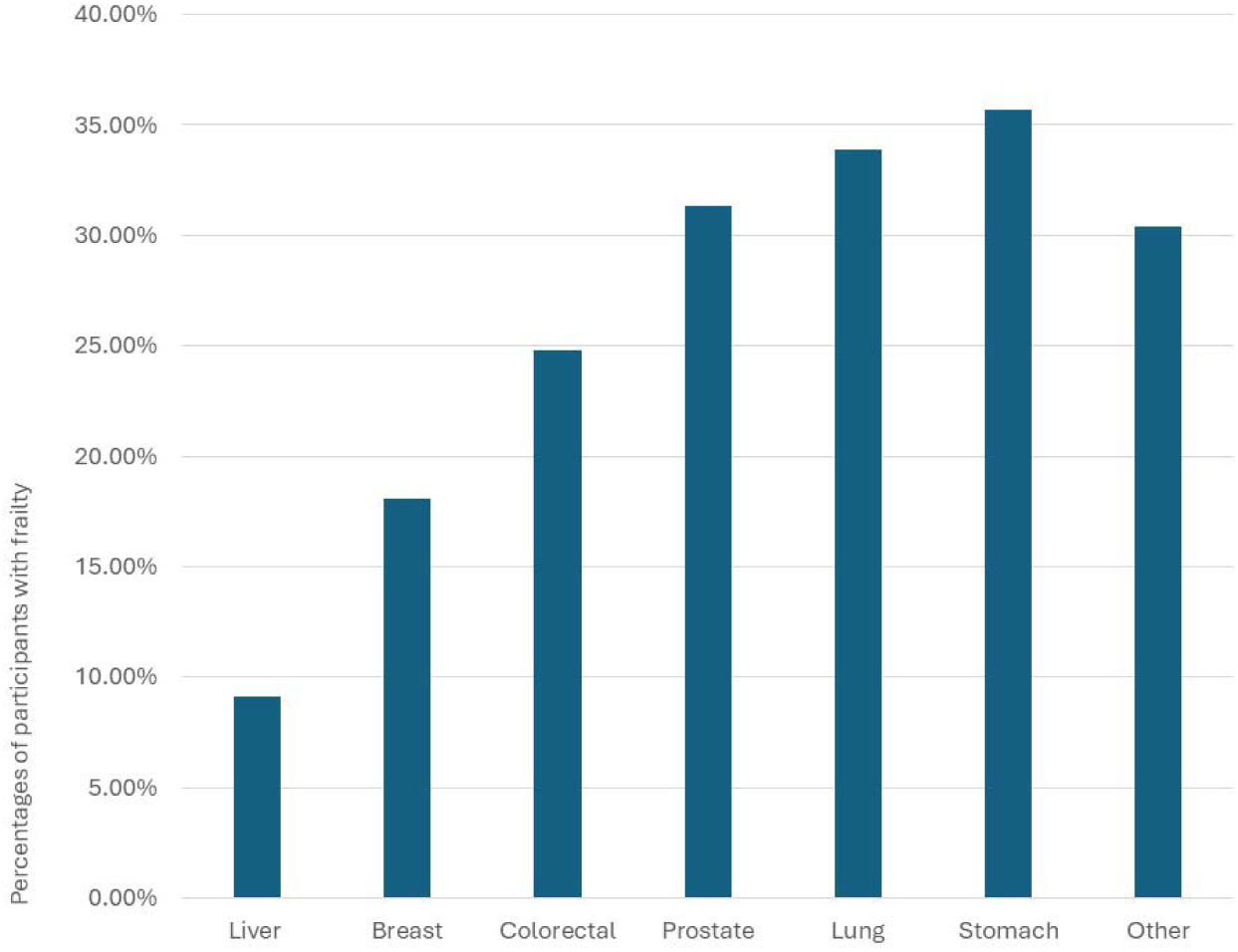
**Frailty prevalence by cancer types**

**Figure 2.**
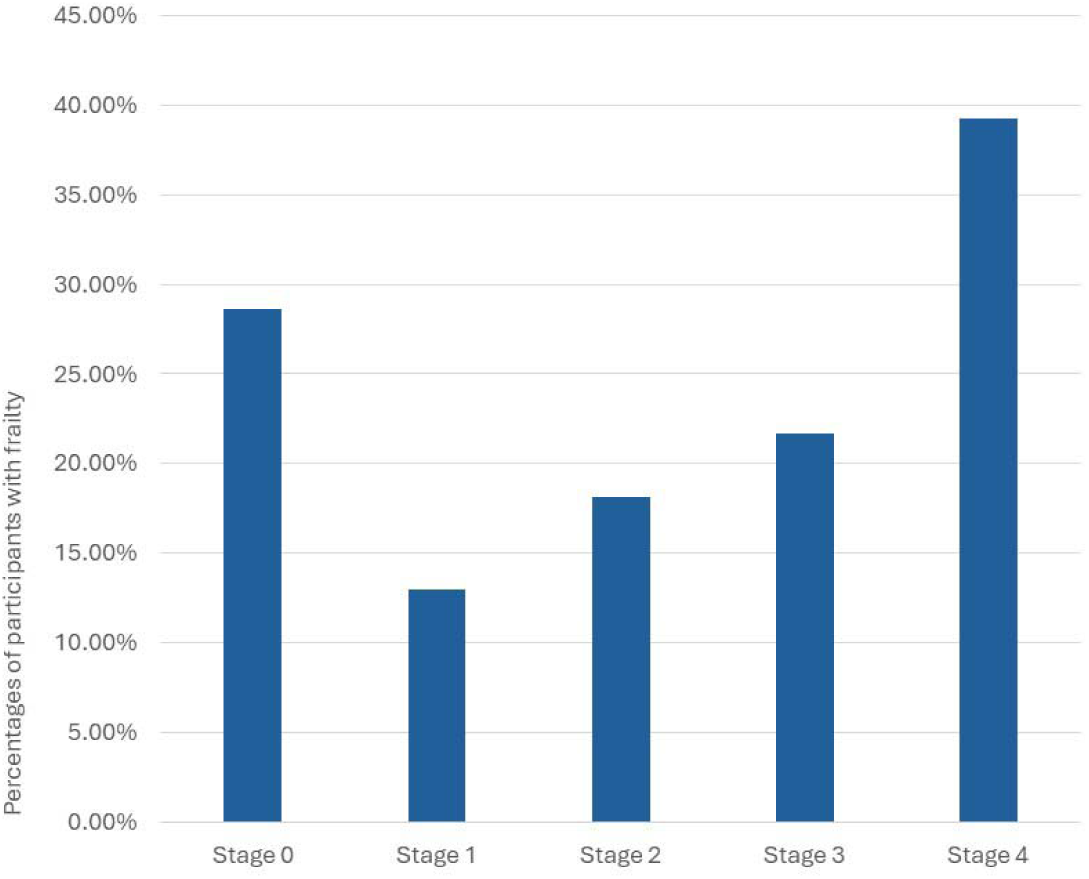
**Frailty prevalence by cancer stages**

Frail participants were significantly older (mean age 75.6 versus 71.1 years in the non-frail, p<0.001). The percentages having a low level of education, being single/ divorced/ widowed, being underweight, and having advanced stages of cancer were higher in frail participants compared to the non-frail. The mean value of the Charlson Comorbidity Index was significantly higher in frail participants (5.0, SD 1.9) compared to non-frail participants (3.7, SD 2.0), p<0.001. The percentages of participants receiving cancer surgery was significantly lower in the frail (57.4%) compared to the non-frail (71.2%, p=0.011), and the proportion of receiving palliative care was significantly higher in the frail (14.9% versus 3.6 in the non-frail, p<0.001) (Table 1 and Table 2)

**Table 1.** Participant characteristics.

| Variables | All participants<br>(N=379) | Non-frail<br>(N=278) | Frail<br>(N=101) | p-values |
| --- | --- | --- | --- | --- |
| Age | 72.3 (6.0) | 71.1 (5.1) | 75.6 (6.9) | <0.001 |
| Age group |  |  |  |  |
| ≤ 70 | 189 (49.9) | 156 (56.1) | 33 (32.7) | <0.001 |
| >70 | 190 (50.1) | 122 (43.9) | 68 (67.3) |  |
| Sex |  |  |  |  |
| Male | 195 (51.5) | 149 (53.6) | 46 (45.5) | 0.166 |
| Female | 184 (48.5) | 129 (46.4) | 55 (54.5) |  |
| Rural/urban living |  |  |  |  |
| Rural | 87 (23.0) | 66 (23.7) | 21 (20.8) | 0.546 |
| Urban | 292 (77.0) | 212 (76.3) | 80 (79.2) |  |
| Marital status |  |  |  |  |
| Married | 284 (74.9) | 219 (78.8) | 65 (64.4) | 0.004 |
| Single/divorced/<br>widowed | 95 (25.1) | 59 (21.2) | 36 (35.6) |  |
| Education |  |  |  |  |
| Primary school | 67 (17.7) | 38 (13.7) | 29 (28.7) | 0.009 |
| Secondary school | 88 (23.2) | 64 (23.0) | 24 (23.8) |  |
| High school | 138 (36.4) | 109 (39.2) | 29 (28.7) |  |
| University/College | 76 (20.1) | 58 (20.9) | 18 (17.8) |  |
| Postgraduate | 10 (2.6) | 9 (3.2) | 1 (1.0) |  |
| Living alone | 26 (6.9) | 17 (6.1) | 9 (8.9) | 0.341 |
| Smoking | 83 (21.9) | 64 (23.0) | 19 (18.8) | 0.381 |
| Regular alcohol consumption | 40 (10.6) | 33 (11.9) | 7 (6.9) | 0.166 |
| Body mass index |  |  |  |  |
| <18.50 | 46 (12.1) | 21 (7.6) | 25 (24.8) | <0.001 |
| 18.50-24.99 | 272 (71.8) | 210 (75.5) | 62 (61.4) |  |
| ≥ 25.0 | 61 (16.1) | 47 (16.9) | 14 (13.9) |  |
| Charlson Comorbidity Index | 4.0 (2.1) | 3.7 (2.0) | 5.0 (1.9) | <0.001 |
| Comorbidities |  |  |  |  |
| Hypertension | 253 (66.8) | 179 (64.4) | 74 (73.3) | 0.105 |
| Coronary heart disease | 95 (25.1) | 70 (25.2) | 25 (24.8) | 0.932 |
| Heart failure | 15 (4.0) | 1 (0.4) | 14 (13.9) | <0.001 |
| Stroke | 8 (2.1) | 3 (1.1) | 5 (5.0) | 0.034 |
| Dyslipidemia | 130 (34.3) | 94 (33.8) | 36 (35.6) | 0.740 |
| Diabetes | 95 (25.1) | 59 (21.2) | 36 (35.6) | 0.004 |
| Chronic kidney disease | 14 (3.7) | 8 (2.9) | 6 (5.9) | 0.162 |
| Osteoarthritis | 59 (15.6) | 36 (12.9) | 23 (22.8) | 0.020 |
| GERD | 53 (14.0) | 35 (12.6) | 18 (17.8) | 0.194 |
| Cognitive decline | 102 (26.9) | 50 (18.0) | 52 (51.5) | <0.001 |
| Anemia | 240 (63.3) | 163 (58.6) | 77 (76.2) | 0.002 |
| Musculoskeletal pain | 202 (53.3) | 134 (48.2) | 68 (67.3) | <0.001 |
| Sleep disorder | 158 (41.7) | 91 (32.7) | 67 (66.3) | <0.001 |
| Anorexia | 125 (33.0) | 66 (23.7) | 59 (58.4) | <0.001 |
| Geriatric syndromes: |  |  |  |  |
| IADL impairment | 214 (56.5) | 119 (42.8) | 95 (94.1) | <0.001 |
| ADL impairment | 6 (1.6) | 1 (0.4) | 5 (5.0) | 0.006 |
| Polypharmacy | 131 (34.6) | 78 (28.1) | 53 (52.5) | <0.001 |
| Malnutrition | 66 (17.4) | 28 (10.1) | 38 (37.6) | <0.01 |
| Depression | 21 (5.5) | 12 (4.3) | 9 (8.9) | 0.084 |
| Having high risk of falls | 161 (42.5) | 78 (28.1) | 83 (82.2) | <0.001 |

Continuous data are presented as mean (standard deviation). Categorical data are shown as n (%). Comparison between frail and non-frail participants were conducted using Chi-square test. GERD: gastroesophageal reflux disease; ADL: Activities of daily living; IADL: instrumental activities of daily living

**Table 2.**
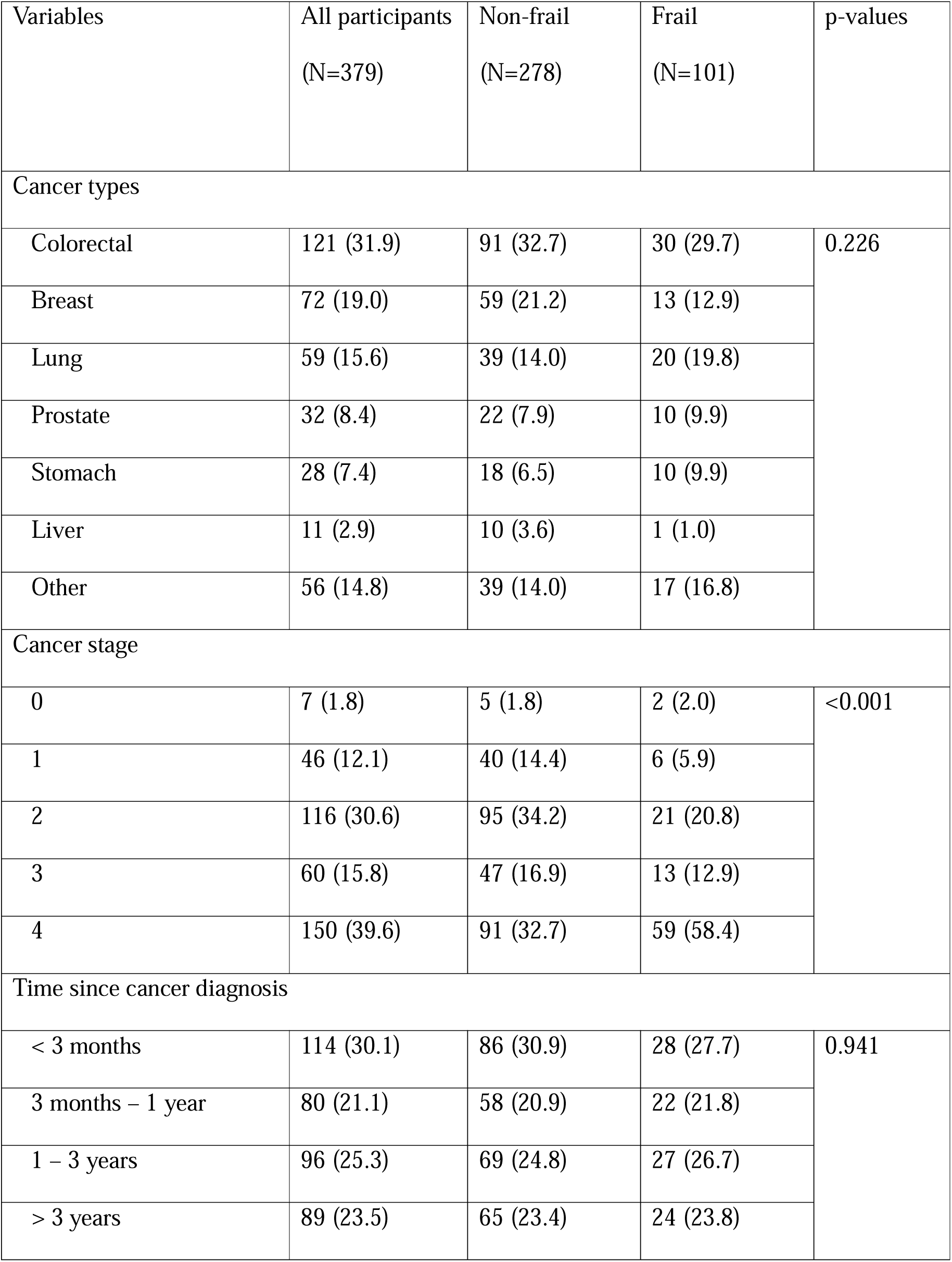
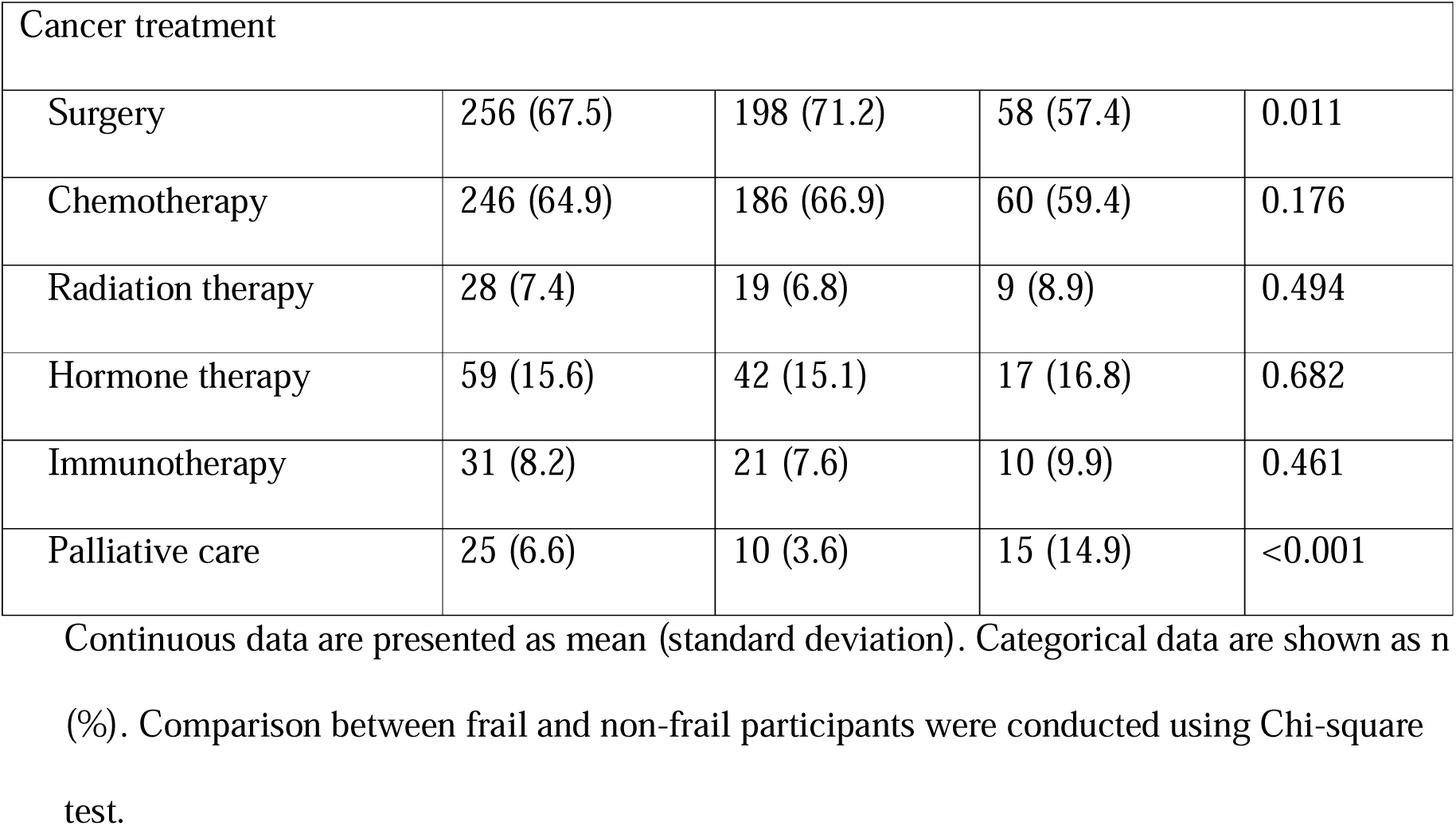
Cancer-related characteristics.

During the 3 months after discharge, 11 participants (2.9%) were lost to follow-up, and data of falls, all-cause hospitalization, all-cause mortality were obtained for 368 participants. The event rates were 19.0% (44.4% in the frail vs. 9.7% in the non-frail, p<0.001) for falls, 33.4% (42.4% in the frail vs. 30.1% in the non-frail, p=0.026) for all-causes hospitalization, and 1.9% (5.1% in the frail vs. 0.7% in the non-frail, p=0.017) for all-cause mortality. (Table 3)

**Table 3.**
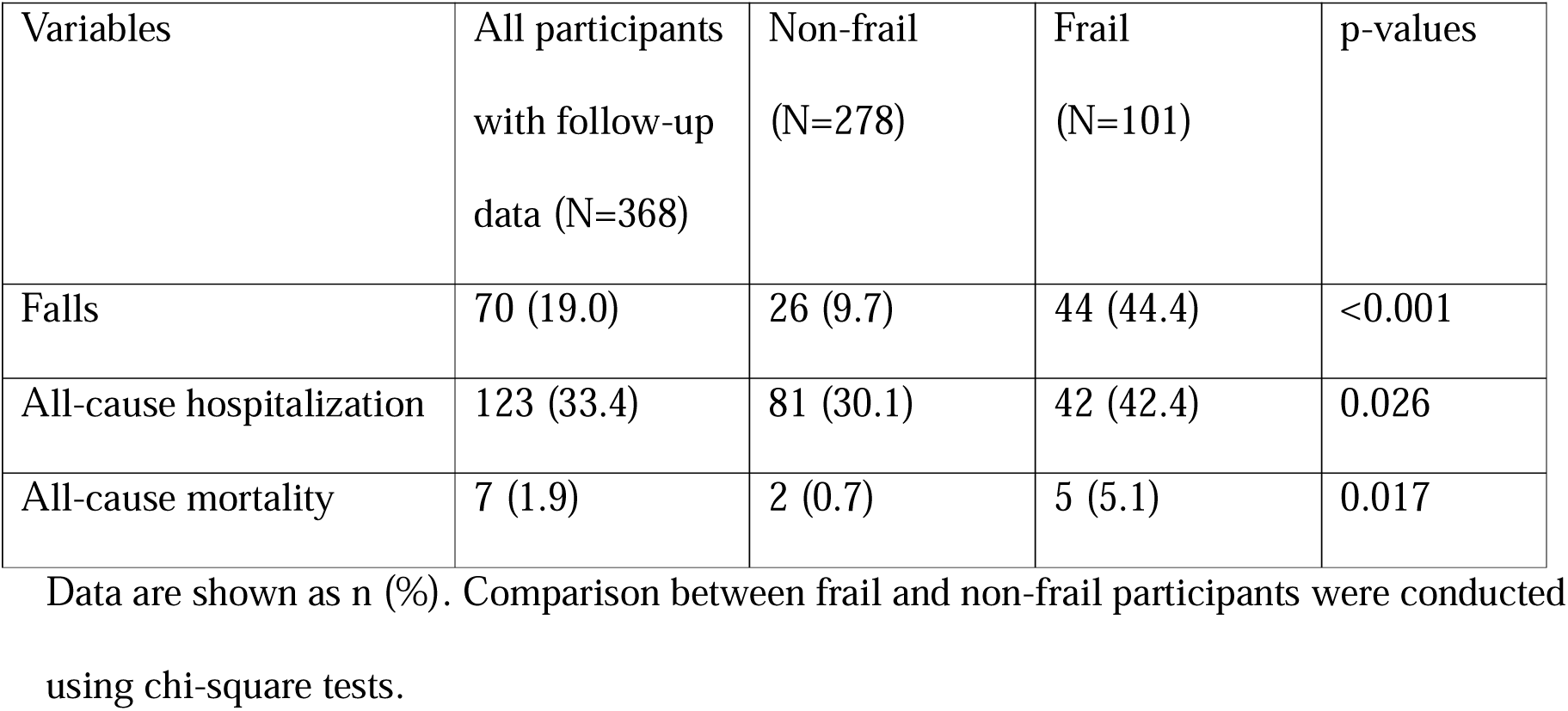
Adverse events by frailty status.

| Variables | All participants<br>with follow-up<br>data (N=368) | Non-frail<br>(N=278) | Frail<br>(N=101) | p-values |
| --- | --- | --- | --- | --- |
| Falls | 70 (19.0) | 26 (9.7) | 44 (44.4) | <0.001 |
| All-cause hospitalization | 123 (33.4) | 81 (30.1) | 42 (42.4) | 0.026 |
| All-cause mortality | 7 (1.9) | 2 (0.7) | 5 (5.1) | 0.017 |

Data are shown as n (%). Comparison between frail and non-frail participants were conducted using chi-square tests.

Frailty was associated with increased odds of falls, all-cause hospitalization and all-cause mortality. The unadjusted ORs were 7.48 (95%CI 4.24 – 13.40, p<0.001) for falls, 1.71 (95%CI 1.06 – 2.75, p=0.027) for all-cause hospitalization, and 7.10 (95%CI 1.36 – 37.22, p=0.020) for all-cause mortality. These relationships are still significant after adjusting for age, sex, and cancer stage (Table 4). Sensitivity analyses showed similar results (Supplementary Tables 1 and 2).

**Table 4.**
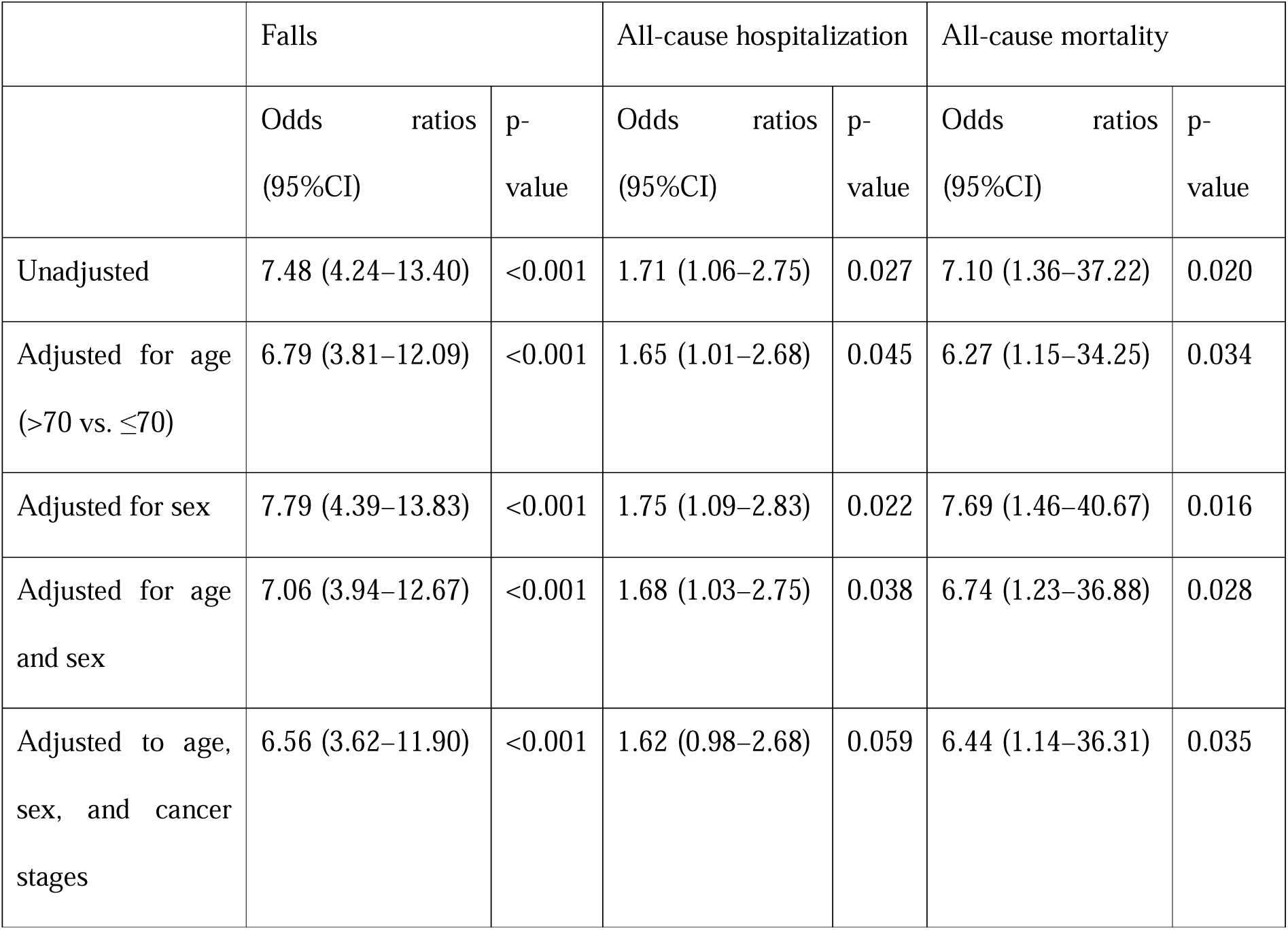
Odds ratios for falls and all-cause hospitalization comparing the frail to the non-frail.

Subgroup analyses showed a non-significant increased impact of frailty on adverse outcomes in men compared to women (Table 5). In models adjusted for age and cancer stage, the ORs were 5.03 (95%CI 2.04 – 12.41) in women vs 7.49 (95%CI 3.37 – 16.67) in men (ROR 0.67, 95%CI 0.20 – 2.24) for falls; 1.22 (95%CI 0.58 – 2.57) in women vs 2.04 (95%CI 1.02 –4.07) in men (ROR 0.60, 95%CI 0.22 – 1.65) for hospitalization; and 2.88 (95%CI 0.12 – 70.38) in women vs 10.42 (95%CI 1.08 – 100.95) in men (ROR 0.28, 95%CI 0.01 – 13.82) for mortality.

**Table 5.** Odds ractios for falls, all-cause hospitalization, and all-cause mortality comparing the frail to the non-frail in women and men.

| Frailty (frail vs. non-frail) | Women (N=184) |  | Men (N=195) |  | Women to men ratio of odds ratios (95%CI) |
| --- | --- | --- | --- | --- | --- |
|  | Odds ratios (95%CI) | p-values | Odds ratios (95%CI) | p-values |  |
| Falls |  |  |  |  |  |
| Unadjusted | 7.48 (3.20 – 17.49) | <0.001 | 8.06 (3.71 – 17.54) | <0.001 | 0.93 (0.29 – 2.93) |
| Adjusted for age (>70 vs ≤70) | 6.97 (2.91 – 16.66) | <0.001 | 7.25 (3.29 – 15.96) | <0.001 | 0.96 (0.30 – 3.12) |
| Adjusted for age (>70 vs ≤70) and cancer stage (≥3 vs ≤2) | 5.03 (2.04 – 12.41) | <0.001 | 7.49 (3.37 – 16.67) | <0.001 | 0.67 (0.20 – 2.24) |
| Hospitalization |  |  |  |  |  |
| Unadjusted | 1.42 (0.72 – 2.82) | 0.314 | 2.15 (1.10 – 4.23) | 0.026 | 0.66 (0.25 – 1.72) |
| Adjusted for age (>70 vs ≤70) | 1.40 (0.69 – 2.83) | 0.354 | 2.03 (1.02 – 4.04) | 0.043 | 0.69 (0.26 – 1.85) |
| Adjusted for age (>70 vs ≤70) and cancer stage (≥3 vs ≤2) | 1.22 (0.58 – 2.57) | 0.608 | 2.04 (1.02 – 4.07) | 0.044 | 0.60 (0.22 – 1.65) |
| Mortality |  |  |  |  |  |
| Unadjusted | 2.37 (0.15 – 38.54) | 0.545 | 13.71 (1.49 – 126.04) | 0.021 | 0.17 (0.00 – 6.03) |
| Adjusted for age (>70 vs ≤70) | 2.62 (0.14 – 47.76) | 0.516 | 11.10 (1.18 – 105.04) | 0.036 | 0.24 (0.01 – 9.36) |
| Adjusted for age (>70 vs ≤70) and cancer stage (≥3 vs ≤2) | 2.88 (0.12 – 70.38) | 0.516 | 10.42 (1.08 – 100.95) | 0.043 | 0.28 (0.01 – 13.82) |

## Discussion

In this study of older patients with cancer, more than a quarter of the participants had frailty, suggesting frailty is a concern for many older patients with cancer and should be considered in the cancer treatment decision-making process. This prevalence accords with studies in older patients with cancer in other Asian countries. For instance, a study conducted in 83 older patients with colorectal cancer who had surgery in Singapore and Japan found that the prevalence of frailty was 28%, using Fried’s frailty phenotype.^26^ Similarly, a study conducted by Yamada and colleagues in 120 older Japanese patients who underwent pancreatic resection for pancreatic ductal adenocarcinoma reported a frailty prevalence of 24.2% (using the Clinical Frailty Scale).^27^ In a study of 662 older patients attending a geriatric oncology clinic in India, 29% were identified as frail, using the Clinical Frailty Scale.^28^ Also, i a Korean study of 391 older adults with cancer living in the community, reported a 24.8% prevalence of frailty, using Fried’s frailty phenotype.^29^ Furthermore, compared with the study conducted by Guerard and colleagues that also used the Carolina Frailty Index to evaluate frailty in older patients with cancer in the US, the prevalence of frailty in our study was slightly higher (26.6% in our study versus 17.8% in theirs), ^25^ which could be due to differences in ethnicity and socioeconomic conditions.

The pathophysiology of frailty involves multiple factors, including chronic systemic inflammation, nutritional deficiencies and impaired neuromuscular function.^30^ In older adults with cancer, especially those in advanced stages, chronic inflammation with increased levels of cytokines such as IL-6, TNF-α and CRP is a significant factor contributing to muscle damage and degeneration.^31–33^ Although the difference in frailty prevalence among participants with different cancer types in this study was not statistically significant, the highest prevalence of frailty was found in participants with stomach cancer (35.7%), followed by lung cancer (33.9%), which is consistent with findings from other countries. A 2024 systematic review of 13 studies worldwide (on 44,117 older adults with gastric cancer) found that the prevalence of frailty was 29% (95% CI 0.21%-0.39%).^34^ A systematic review (published in 2022) of 16 articles involving 4,183 patients with lung cancer worldwide found that the prevalence of frailty was 45% (95% CI 28%-61%).^35^ The exact reason why patients with stomach and lung cancer had the highest prevalence of frailty is not well known. But the possible justification could be that patients with lung cancer have decreased respiratory function and severely limited mobility. Common symptoms of lung cancer, such as shortness of breath and chronic fatigue, often lead to reduced physical activity, thereby accelerating the process of muscle mass loss and reduced motor function.^35,36^ Stomach cancer can lead to malnutrition and reduced ability to absorb nutrients, contributing to the development of frailty.^33,37^

We found that the prevalence of frailty was higher in patients with advance stages of cancer, which is similar to findings from Guerard and colleagues in 546 patients with cancer (median age 72 years) in the Carolina Senior UNC Registry for Older Patients.^25^ Patients with advanced cancer often undergo intense treatments such as chemotherapy or radiotherapy, which contribute to an increased risk of malnutrition and severe muscle waste.

Frailty in patient with cancer is associated with an increased risk of adverse outcomes, such as postoperative complications, treatment intolerance, disease progression, increased hospitalization, and mortality.^38,39^ These highlight the crucial role of frailty assessment in the management of older patients with cancer. Our study found that frailty is strongly associated with falls, all-cause hospitalization and all-cause mortality in the three months of follow-up. Musculoskeletal weakness is a significant component of the frailty syndrome. In older patients with cancer, the impact of cancer treatment and its side effects, including anorexia and malnutrition, can lead to a decline in both muscle mass and strength. This condition not only limits mobility but can also impairs ability to perform daily activities, and increases the risk of falls and fractures.^40–42^ Previous studies in other countries showed that frailty status was associated with increased hospital admissions. Williams et al. evaluated the associations between frailty and long-term functional outcomes and found that frailty increased the risk of hospital admission by 2.5-fold and long-term care admission by 1.9-fold.^43^ Shahrokni et al., in a cohort of 1137 patients with cancer, reported that higher frailty scores were significantly associated with a longer postsurgical length of stay and a higher risk of intensive care unit admission and 1-year mortality.^44^ In a 2015 systematic review including 20 observational studies (2916 participants) with data on the prevalence and/or outcomes of frailty in older cancer patients, frailty was independently associated with increased all-cause mortality, and increased intolerance to cancer treatment.^3^ Another review on the impact of frailty on health outcomes in older adults with lung cancer, revealed that frailty had a strong and consistent association with mortality.^45^

Our findings have important clinical implications. Understanding the burden of frailty in older patients with cancer is crucial because it significantly impacts treatment outcomes and quality of life. Frailty measures the overall health and ability to recover, which can vary widely among older adults facing cancer. Understanding frailty helps healthcare providers tailor treatments to individual needs, avoiding overly aggressive therapies that may be harmful or less effective. It also aids in predicting potential complications and guiding decisions around palliative care, ensuring a balanced approach that prioritizes both survival and well-being. It is crucial to implement frailty screening programs and assess frailty regularly during cancer treatment. Integrating frailty assessment into cancer care fosters more personalized, compassionate treatment plans, enhancing patient outcomes and quality of life. In addition, by identifying frailty early on, interventions can be implemented timely to reduce the risk of complications and adverse outcomes. Frail and pre-frail patients should be regularly monitored for falls risk.

To the best of our understanding, this is the first study in Vietnam examining frailty and its impact on adverse outcomes in older patients with cancer, using a validated frailty assessment tool. However, the study was conducted at two urban hospitals in one city, which may not be representative for all older patients with cancer in Vietnam. Therefore, the findings should be interpreted with caution when considering their application to different healthcare systems or policy contexts. In addition, the follow up duration was short, and other important outcomes for patients with cancer, such as quality of life, were not examined.

## Conclusions

In this study of older patients with cancer, frailty was observed in over a quarter of the participants, with the highest prevalence of frailty found in participants with stomach cancer and lung cancer. Frailty significantly increased the odds of falls, hospitalization, and mortality by three months post-discharge. Further research with a larger sample size and longer follow up is necessary to gain a better understanding of the impact of frailty on adverse outcomes, as well as quality of life in older adults with cancer in Vietnam, and whether there is any sex difference in the impact of frailty. Future investigations on frailty in older patients with cancer should also focus on its impact on cancer treatment decisions and outcomes.

## Data Availability

All data produced in the present study are available upon reasonable request to the authors.

## References

1. Siegel RL, Miller KD, Jemal A. Cancer Statistics, 2017. CA Cancer J Clin 2017; 67(1): 7-30.

2. Ethun CG, Bilen MA, Jani AB, Maithel SK, Ogan K, Master VA. Frailty and cancer: Implications for oncology surgery, medical oncology, and radiation oncology. CA Cancer J Clin 2017; 67(5): 362–77.

3. Handforth C, Clegg A, Young C, et al. The prevalence and outcomes of frailty in older cancer patients: a systematic review. Annals of Oncology 2015; 26(6): 1091–101.

4. Fletcher JA, Fox ST, Reid N, Hubbard RE, Ladwa R. The impact of frailty on health outcomes in older adults with lung cancer: A systematic review. Cancer Treat Res Commun 2022; 33: 100652.

5. Overcash J, Cope DG, Van Cleave JH. Frailty in Older Adults: Assessment, Support, and Treatment Implications in Patients With Cancer. Clin J Oncol Nurs 2018; 22(6): 8–18.

6. Nguyen SM, Deppen S, Nguyen GH, Pham DX, Bui TD, Tran TV. Projecting Cancer Incidence for 2025 in the 2 Largest Populated Cities in Vietnam. Cancer Control 2019; 26(1): 1073274819865274.

7. The ASG. Catastrophic health expenditure and 12-month mortality associated with cancer in Southeast Asia: results from a longitudinal study in eight countries. BMC Medicine 2015; 13(1): 190.

8. Nguyen TV, Tran HM, Trinh HBT, Vu VH, Bang VA. Prevalence of frailty according to the Hospital Frailty Risk Score and related factors in older patients with acute coronary syndromes in Vietnam. Australas J Ageing 2024.

9. Nguyen AT, Nguyen TX, Nguyen TN, et al. The impact of frailty on prolonged hospitalization and mortality in elderly inpatients in Vietnam: a comparison between the frailty phenotype and the Reported Edmonton Frail Scale. Clin Interv Aging 2019; 14: 381–8.

10. Nguyen HT, Do HT, Nguyen HVB, Nguyen TV. Fried Frailty Phenotype in Elderly Patients with Chronic Coronary Syndrome: Prevalence, Associated Factors, and Impact on Hospitalization. J Multidiscip Healthc 2024; 17: 1265–74.

11. Nguyen AT, Nguyen LH, Nguyen TX, et al. Frailty Prevalence and Association with Health-Related Quality of Life Impairment among Rural Community-Dwelling Older Adults in Vietnam. Int J Environ Res Public Health 2019; 16(20).

12. Nguyen TV, Le D, Tran KD, Bui KX, Nguyen TN. Frailty in Older Patients with Acute Coronary Syndrome in Vietnam. Clin Interv Aging 2019; 14: 2213–22.

13. Nguyen HT, Nguyen AH, Le PTM. Sex differences in frailty of geriatric outpatients with type 2 diabetes mellitus: a multicentre cross-sectional study. Scientific Reports 2022; 12(1): 16122.

14. Khuc AHT, Doan VT, Le TT, et al. Determinants of Frailty among Patients with Type 2 Diabetes In Urban Hospital. Hosp Top 2023; 101(3): 215-22.

15. Huynh TQH, Pham TLA, Vo VT, Than HNT, Nguyen TV. Frailty and Associated Factors among the Elderly in Vietnam: A Cross-Sectional Study. Geriatrics (Basel) 2022; 7(4).

16. Vu HTT, Nguyen TX, Nguyen TN, et al. Prevalence of frailty and its associated factors in older hospitalised patients in Vietnam. BMC Geriatr 2017; 17(1): 216.

17. Masnoon N, Shakib S, Kalisch-Ellett L, Caughey GE. What is polypharmacy? A systematic review of definitions. BMC Geriatr 2017; 17(1): 230.

18. Tuch G, Soo WK, Luo KY, et al. Cognitive Assessment Tools Recommended in Geriatric Oncology Guidelines: A Rapid Review. Curr Oncol 2021; 28(5): 3987–4003.

19. Gómez JM, Carrera F. What should the optimal target hemoglobin be? Kidney Int Suppl 2002; (80): 39–43.

20. Hoyl MT, Alessi CA, Harker JO, et al. Development and testing of a five-item version of the Geriatric Depression Scale. J Am Geriatr Soc 1999; 47(7): 873–8.

21. Charlson ME, Pompei P, Ales KL, MacKenzie CR. A new method of classifying prognostic comorbidity in longitudinal studies: Development and validation. Journal of Chronic Diseases 1987; 40(5): 373–83.

22. Kaiser MJ, Bauer JM, Ramsch C, et al. Validation of the Mini Nutritional Assessment short-form (MNA-SF): a practical tool for identification of nutritional status. J Nutr Health Aging 2009; 13(9): 782–8.

23. Lin CC, Meardon S, O’Brien K. The Predictive Validity and Clinical Application of Stopping Elderly Accidents, Deaths & Injuries (STEADI) for Fall Risk Screening. Adv Geriatr Med Res 2022; 4(3).

24. Katz S. Assessing self-maintenance: activities of daily living, mobility, and instrumental activities of daily living. J Am Geriatr Soc 1983; 31(12): 721–7.

25. Guerard EJ, Deal AM, Chang Y, et al. Frailty Index Developed From a Cancer-Specific Geriatric Assessment and the Association With Mortality Among Older Adults With Cancer. J Natl Compr Canc Netw 2017; 15(7): 894–902.

26. Tan KY, Kawamura YJ, Tokomitsu A, Tang T. Assessment for frailty is useful for predicting morbidity in elderly patients undergoing colorectal cancer resection whose comorbidities are already optimized. Am J Surg 2012; 204(2): 139–43.

27. Yamada S, Shimada M, Morine Y, et al. Significance of frailty in prognosis after surgery in patients with pancreatic ductal adenocarcinoma. World J Surg Oncol 2021; 19(1): 94.

28. Rao AR, Ramaswamy A, Kumar S, et al. Prevalence and outcomes of frailty in older patients with cancer: A prospective study from geriatric oncology clinic. Journal of Clinical Oncology; 40(16_suppl): e24011-e.

29. Kang H. Correlates of Frailty in Community-Dwelling Older Adults with Cancer: 2017 Survey of Living Condition of Elderly Study in South Korea. Asia Pac J Oncol Nurs 2021; 8(3): 287–94.

30. Clegg A, Young J, Iliffe S, Rikkert MO, Rockwood K. Frailty in elderly people. The Lancet 2013; 381(9868): 752-62.

31. Lan T, Chen L, Wei X. Inflammatory Cytokines in Cancer: Comprehensive Understanding and Clinical Progress in Gene Therapy. Cells 2021; 10(1).

32. Schmidt SF, Rohm M, Herzig S, Berriel Diaz M. Cancer Cachexia: More Than Skeletal Muscle Wasting. Trends Cancer 2018; 4(12): 849–60.

33. Aversa Z, Costelli P, Muscaritoli M. Cancer-induced muscle wasting: latest findings in prevention and treatment. Ther Adv Med Oncol 2017; 9(5): 369–82.

34. Tang WZ, Tan ZK, Qiu LY, Chen JQ, Jia K. Prevalence and unfavorable outcome of frailty in older adults with gastric cancer: a systematic review and meta-analysis. Support Care Cancer 2024; 32(2): 115.

35. Komici K, Bencivenga L, Navani N, et al. Frailty in Patients With Lung Cancer: A Systematic Review and Meta-Analysis. Chest 2022; 162(2): 485–97.

36. Venuta F, Diso D, Onorati I, Anile M, Mantovani S, Rendina EA. Lung cancer in elderly patients. J Thorac Dis 2016; 8(Suppl 11): S908–s14.

37. Zheng J, Wang X, Yu J, et al. Global Leadership Initiative on Malnutrition criteria: Clinical benefits for patients with gastric cancer. Nutr Clin Pract 2025; 40(1): 239–51.

38. Ethun CG, Bilen MA, Jani AB, Maithel SK, Ogan K, Master VA. Frailty and cancer: implications for oncology surgery, medical oncology, and radiation oncology. CA: a cancer journal for clinicians 2017; **67**(5): 362-77.

39. Ness KK, Wogksch MD. Frailty and aging in cancer survivors. Translational Research 2020; 221: 65–82.

40. Dunne RF, Loh KP, Williams GR, Jatoi A, Mustian KM, Mohile SG. Cachexia and Sarcopenia in Older Adults with Cancer: A Comprehensive Review. Cancers (Basel) 2019; 11(12).

41. Ali S, Garcia JM. Sarcopenia, cachexia and aging: diagnosis, mechanisms and therapeutic options - a mini-review. Gerontology 2014; 60(4): 294–305.

42. Morris R, Lewis A. Falls and Cancer. Clinical Oncology 2020; 32(9): 569–78.

43. Williams GR, Dunham L, Chang Y, et al. Geriatric assessment predicts hospitalization frequency and long-term care use in older adult cancer survivors. Journal of oncology practice 2019; 15(5): e399–e409.

44. Shahrokni A, Tin A, Alexander K, et al. Development and evaluation of a new frailty index for older surgical patients with cancer. JAMA Network Open 2019; 2(5): e193545-e.

45. Fletcher JA, Fox ST, Reid N, Hubbard RE, Ladwa R. The impact of frailty on health outcomes in older adults with lung cancer: A systematic review. Cancer treatment and research communications 2022; 33: 100652.

